# COVID Oximetry @home: evaluation of patient outcomes

**DOI:** 10.1101/2021.05.29.21257899

**Authors:** Michael Boniface, Daniel Burns, Chris Duckworth, Mazen Ahmed, Franklin Duruiheoma, Htwe Armitage, Naomi Ratcliffe, John Duffy, Caroline O’Keeffe, Matt Inada-Kim

## Abstract

**Background:** COVID-19 has placed unprecedented demands on hospitals. A clinical service, COVID Oximetry @home (CO@h) was launched in November 2020 to support remote monitoring of COVID-19 patients in the community. Remote monitoring through CO@h aims to identify early patient deterioration and provide timely escalation for cases of silent hypoxia, while reducing the burden on secondary care.

**Methods:** We conducted a retrospective service evaluation of COVID-19 patients onboarded to CO@h from November 2020 to March 2021 in the North Hampshire (UK) community led service (a collaboration of 15 GP practices covering 230,000 people). We have compared outcomes for patients admitted to Basingstoke & North Hampshire Hospital who were CO@h patients (COVID-19 patients with home monitoring of SpO_2_ (n=115)), with non-CO@h patients (those directly admitted without being monitored by CO@h (n=633)). Crude and adjusted odds ratio analysis was performed to evaluate the effects of CO@h on patient outcomes of 30-day mortality, ICU admission and hospital length of stay greater than 3, 7, 14, and 28 days.

**Results:** Adjusted odds ratios for CO@h show an association with a reduction for several adverse patient outcome: 30-day hospital mortality (p<0.001 OR 0.21 95% CI 0.08-0.47), hospital length of stay larger than 3 days (p<0.05, OR 0.62 95% CI 0.39-1.00), 7 days (p<0.001 OR 0.35 95% CI 0.22-0.54), 14 days (p<0.001 OR 0.22 95% CI 0.11-0.41), and 28 days (p<0.05 OR 0.21 95% CI 0.05-0.59). No significant reduction ICU admission was observed (p>0.05 OR 0.43 95% CI 0.15-1.04). Within 30 days of hospital admission, there were no hospital readmissions for those on the CO@h service as opposed to 8.7% readmissions for those not on the service.

**Conclusions:** We have demonstrated a significant association between CO@h and better patient outcomes; most notably a reduction in the odds of hospital lengths of stays longer than 7, 14 and 28 days and 30-day hospital mortality.

## Introduction

The rapidly evolving COVID-19 pandemic has been responsible for 3.4 million deaths worldwide (World Health Organization, 2021) and has placed unprecedented strain on healthcare systems. A significant proportion of patients hospitalised with acute COVID-19 have severe hypoxia (very low blood oxygen saturation) frequently presenting ‘silently’ (i.e. without breathlessness) (Greenhalgh, et al., 2021). Silent hypoxia is an independent indicator of worse patient outcomes (O’Carroll, et al., 2020) (Brouqui, et al., 2021), and delayed presentations of severe COVID-19; often leading to extended hospital stays, higher risk of ICU admission, and higher mortality rates (Vindrola-Padros, et al., 2020).

UK guidelines recommend that patient acuity should therefore be assessed with the use of pulse oximetry (i.e. monitoring oxygen saturation) when diagnosed with COVID-19 (NHS England, 2020) (Inada-Kim, et al., 2020). A clinical service COVID Oximetry @home (CO@h) was launched November 2020 within a COVID-19 Integrated Care Pathway to support remote monitoring of COVID-19 patients by primary care and timely escalation to secondary care. Remote home monitoring through CO@h have been implemented to 1) maintain NHS capacity, 2) decrease nosocomial COVID-19 transmission, and 3) identify early patient deterioration and provide timely escalation to reduce hospital length of stay, and mortality from silent hypoxia (Stockly, 2020) (Boniface, Zlatev, Guerrero-Luduena, & Armitage, 2020).

A CO@h Service consists of two fundamental components: (1) using a predictive model to identify individual patients in a population who are at high risk of future unplanned hospital admission; and (2) offering these people a period of intensive, multidisciplinary, case management at home using the systems, staffing and daily routine (Lewis et al., 2006) (Lewis, et al., 2013). Patients are referred by clinical services responsible for operating CO@h services then triaged prior to onboarding for remote monitoring to ensure that the CO@h offers an appropriate level of care.

Even though the CO@h service is virtual, it is integrated into a care partnership with physical care for patient assessment (e.g. COVID-19 testing and initial observations) when deemed clinically appropriate. A schematic of the Integrated Care Partnership for North Hampshire is shown in Figure 1, highlighting the important relationship between physical and virtual services in the overall delivery of care.

**Figure 1:**
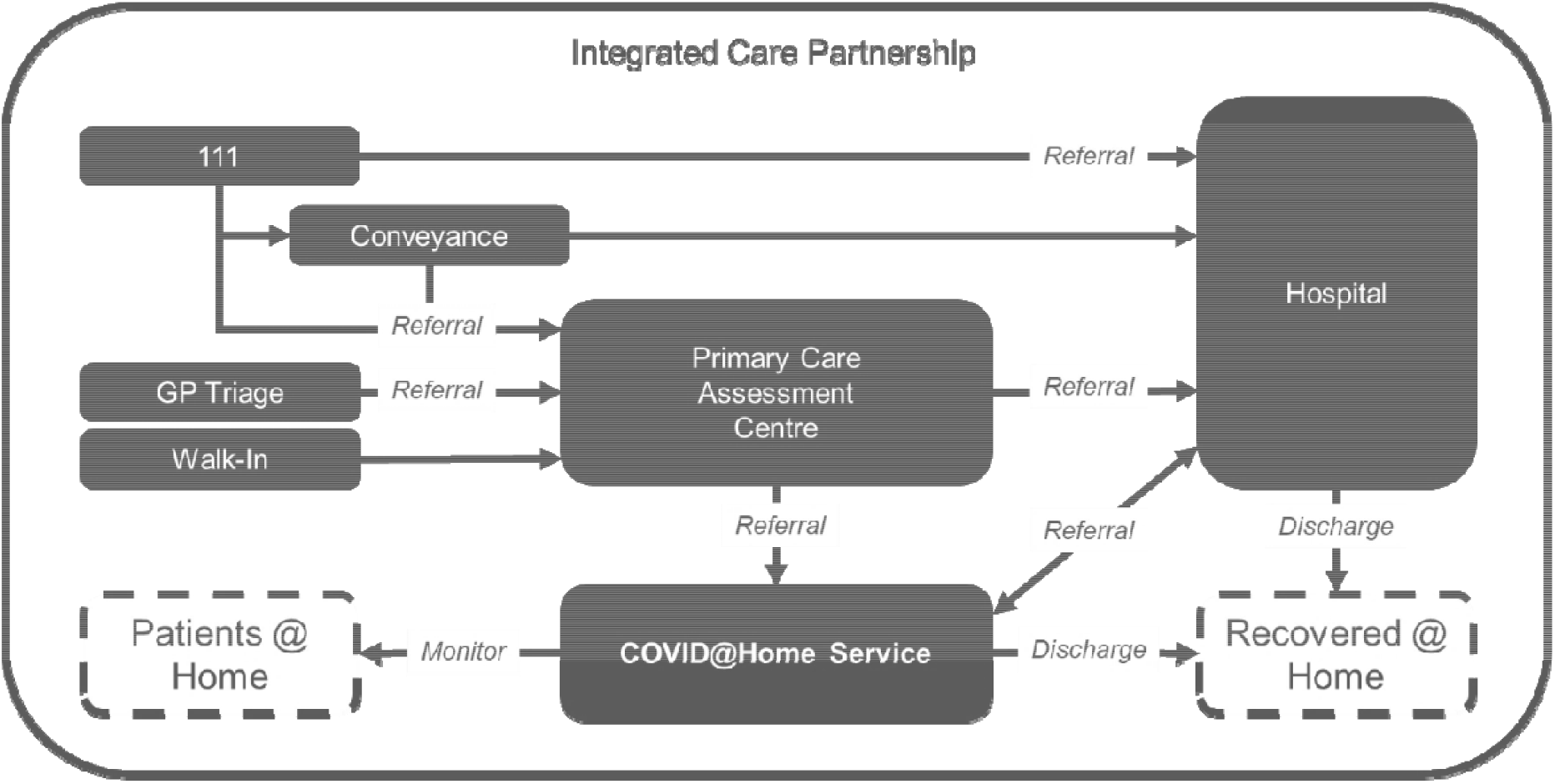
Primary care assessment centre and COVID Oximetry @home service deployed supporting community patient monitoring as defined by North Hampshire Integrated Care System.

This report describes a quality improvement (QI) initiative to implement CO@h service in North Hampshire and retrospectively evaluate their efficacy. Through adoption of the Plan-Do-Study-Act (PDSA) framework and using evidence-based practice, the CO@h services were continually improved as a rapid response to the evolving pandemic. Evidence-based practice was enabled by close communication between professional analysts, who provided data insight, and healthcare professionals, who provided operational insight.

North Hampshire covers 230,000 patients across a single Clinical Commissioning Group and six Primary Care Networks served by one acute hospital trust. The North Hampshire COVID Oximetry@Home Standard Operating Procedure v7.0 was approved October 2020 with service going live November 2020. Patients accessing CO@h were assessed face to face to determine those at high risk of deterioration. The assessment included a COVID-19 swab test, baseline measurements (Pulse, SpO_2_ and NEWS2) and risk factors (ethnicity, age, BMI, comorbidities, mental health, shielding, and high-risk professions). Only patients with a SARS-CoV-2 positive test were considered for subsequent pathways categorised by severity of red, amber and green. Red patients with SpO_2_ ≤ 92% or any of respiration rate ≥ 25, heart rate ≥ 131, new confusion and NEWS2 ≥ 5 were admitted directly to hospital. Amber patients with 93% ≤ SpO_2_ ≤ 94%, or any of 21 ≤ respiration rate ≤ 24, 91 ≤ heart rate ≤ 130, 3 ≤ NEWS2 ≤ 4 had further tests (chest x-ray, blood and desaturation) to determine if hospital admission is needed for drug therapeutics (Remdesivir or Dexamethasone) or if not then admitted to CO@h. Green patients identified with risk factors and SpO_2_ ≥ 95%, or any of respiration rate ≤ 20, heart rate ≤ 90, 0 ≤ NEWS2 ≤ 2 were admitted to CO@h.

All patients admitted to CO@h were issued with a standard pulse oximeter and information sheet describing how to use a pulse oximeter and the protocol for reporting SpO_2_ measurements three times a day using either a remote monitoring mobile application or paper diaries. Patients were remotely monitored for up to 14 days, and escalation decisions made in accordance with red, amber and green criteria described in the categories above.

It should be noted that pilot services were operational in North Hampshire during the 1st wave of the pandemic (April 2020 to June 2020) to iteratively explore the definition of operating procedures. Data relating to CO@h pilots during the 1st wave period was not available for analysis. In this report we explore the effectiveness of the CO@h service implemented in the Integrated Care Partnership of North Hampshire. To our knowledge, this is the first QI initiative directly reporting outcomes for COVID-19 patients treated virtually for an NHS Trust. Our findings will be of interest to healthcare organisations looking to implement further CO@h services as a response to the ongoing pandemic.

## Methods

### Patient Population

All patients with suspected COVID-19 admitted to North Hants CO@h or Basingstoke and North Hants Hospital (BNHH) between 1 November 2020 to 31 March 2021 were eligible for inclusion. CO@h patients were then linked to their subsequent hospital admissions. Confirmed COVID-19 cases were then identified from these suspected cases by requiring at least one SARS-CoV-2 positive test associated with the admission.

We separated our cohort into an intervention group, where patients had at least one referral to the CO@h service, and a control group, where patients have not had such a referral. There was a total of 1496 patients, with 783 patients in the intervention group and 713 patients in the control group. To evaluate the outcomes between comparable groups, we required that each patient in the intervention group had at least one hospital admission via the emergency department. We excluded patients in the intervention group who returned a negative COVID test result following referral to CO@h (n=35) and those whose outcomes did not result in escalation to hospital (n=611). We also excluded patients who did not engage or had a CO@h admission date after hospital admission (n=22), leaving 115 CO@h patients that were escalated to hospital in our intervention group.

For the control group we excluded COVID-19 patients (n=80) admitted from hospital locations other than the emergency department to reduce confounding factors between the intervention and control groups: patients in this group may have already been admitted to hospital with complex ongoing acute care needs in addition to COVID-19; resulting in an increased likelihood for negative outcomes, such as longer length of stay and hospital mortality. This left 633 patients in the Non-CO@h control group, of which 55 patients were readmitted to hospital within 30 days of first admission. For readmissions an episode of care was created by aggregating the length of stay over all admissions. Patient outcome was then deemed to be the outcome from the last admission event.

### Target Outcome

We evaluated the CO@h service using a comparison of increasingly acute outcomes associated with patient trajectory for those with a positive COVID-19 test who required admission to hospital. We considered the following outcomes: 30-day hospital mortality, ICU admission, and hospital length of stay above 3 days, 7 days, 14 days, or 28 days. We identified mortality through hospital medical records. We identified ICU admissions through a specific electronic patient record flag. The length of stay was computed from the point of hospital admission to discharge.

### Data Collection

We linked data from Primary Care systems operated by CO@h with Secondary Care systems operated by Hampshire Hospitals NHS Foundation Trust (HHFT) to create a database supporting analysis of the full trajectory of COVID-19 episodes. The linking included CO@h service and hospital admission records.

To ensure data quality, we subsequently excluded admission records in the overall patient polutation that did not meet the following criteria: (1) admission date must be equal to or before the discharge date; (2) the discharge date and patient outcome must be recorded. There were 2 hospital admission records with a discharge date before the corresponding admission date, and 11 admissions without a discharge date or outcome. There were 2 instances of duplicates: 1 duplicate set of CO@h referral records, and 1 duplicate set of hospital admission records that were removed.

The dataset also contains some hospital admission records for which the date of admission is prior to discharge from a CO@h service. In some cases, patients admitted to hospital were not discharged from CO@h immediately. If the hospital admission was likely to only be less than 24hrs (i.e., same day emergency care), patients would continue intervention by CO@h seamlessly as they left hospital. If hospital admission was longer than 24 hours, patients were discharged from CO@h, and in some cases referred back to CO@h following hospital discharge. Patients were also given advice to contact 999, or another service based on their self-submitted readings. In some cases, the patient would then be conveyed and admitted to hospital ahead of discharge from the CO@h service. In all cases, we include those patients within our intervention group as they have received the CO@h intervention.

Confounding comorbidity risks were included for patient conditions that increased the chance of a negative outcome from COVID-19 (Williamson, et al., 2020) (Dengl, Yin, Chen, & Zeng, 2020) (Docherty, et al., 2020) (Sanyaolu, et al., 2020). These conditions include: cardiac disease, diabetes, pulmonary disease, asthma, obesity, respiratory disease, chronic heart disease, liver disease, stroke, dementia, autoimmune disease, malignancy, dialysis, renal failure, cardiac disease, kidney disease, kidney failure, cardiovascular disease, hypertension, cancer, hyperlipidemia, coronary artery disease, renal disease, COPD, atrial fibrillation and heart failure.

### Data Analysis

A time series was produced to show the monthly CO@h referrals with number of hospital admissions from November 2020 to March 2021. An empirical cumulative distribution function was computed for length of stay in the intervention and control groups, in addition to their smoothed distribution functions (calculated by kernel density estimate with a Gaussian kernel restricted to positive values).

Pearson’s chi squared test was performed to check for a significant difference in distributions of patient characteristics in the intervention and control group, where p<0.05 is considered significant. We used logistic regression to account for possible confounders for each patient outcome and calculated the crude odds ratio of CO@h and adjusted odds ratios for patient’s age group, gender and comorbidities. We utilise the Wald test to evaluate whether logistic regression coefficients are significant with respect to the null hypothesis that the resulting odds ratios are 1. Analysis was performed in Python v3.9.4 (using pandas, seaborn, statsmodels) and R 3.6.1.

### Data Governance

This service evaluation did not require ethics approval. The study was however evaluated by the University of Southampton Ethics Committee (REF ERGO/61242) and approved as a service evaluation following Data Protection Impact Assessment and establishment of Data Sharing Agreements. NHS England and NHS Improvement have been given legal notice by the Secretary of State for Health and Social Care to support the processing and sharing of information to help the COVID-19 response under Health Service Control of Patient Information Regulations 2002 (COPI). This is to ensure that confidential patient information can be used and shared appropriately and lawfully for purposes related to the COVID-19 response. Data were extracted from medical records by clinicians providing care for the patients and an anonymised extract of the data were provided to the team at the University of Southampton.

### Data Accessibility

Due to information governance concerns, the data will not be made public. However, it may be made accessible via reasonable request to the corresponding author.

## Results

The North Hants CO@h service treated 783 patients of which 115 patients were subsequently escalated and admitted to hospital. The Basingstoke and North Hants hospital had a further 692 admissions (633 individual patients) directly via the emergency department who were not treated by the CO@h service.

The uptake of the CO@h service is shown in Figure 3. The ratio of CO@h referrals to the number of hospital admissions increased consistently throughout the period, with a ratio of 30/113 (1:4) during November 2020, to 85/21 (4:1) during March 2021. The uptake reached a maximum of 363 referrals during January 2021 coinciding with the increase in COVID-19 prevalence during that period. The mean length of stay for CO@h patients who were then subsequently escalated to hospital was 11.62 days [95% CI 10.76 – 12.48 days].

**Figure 2:**
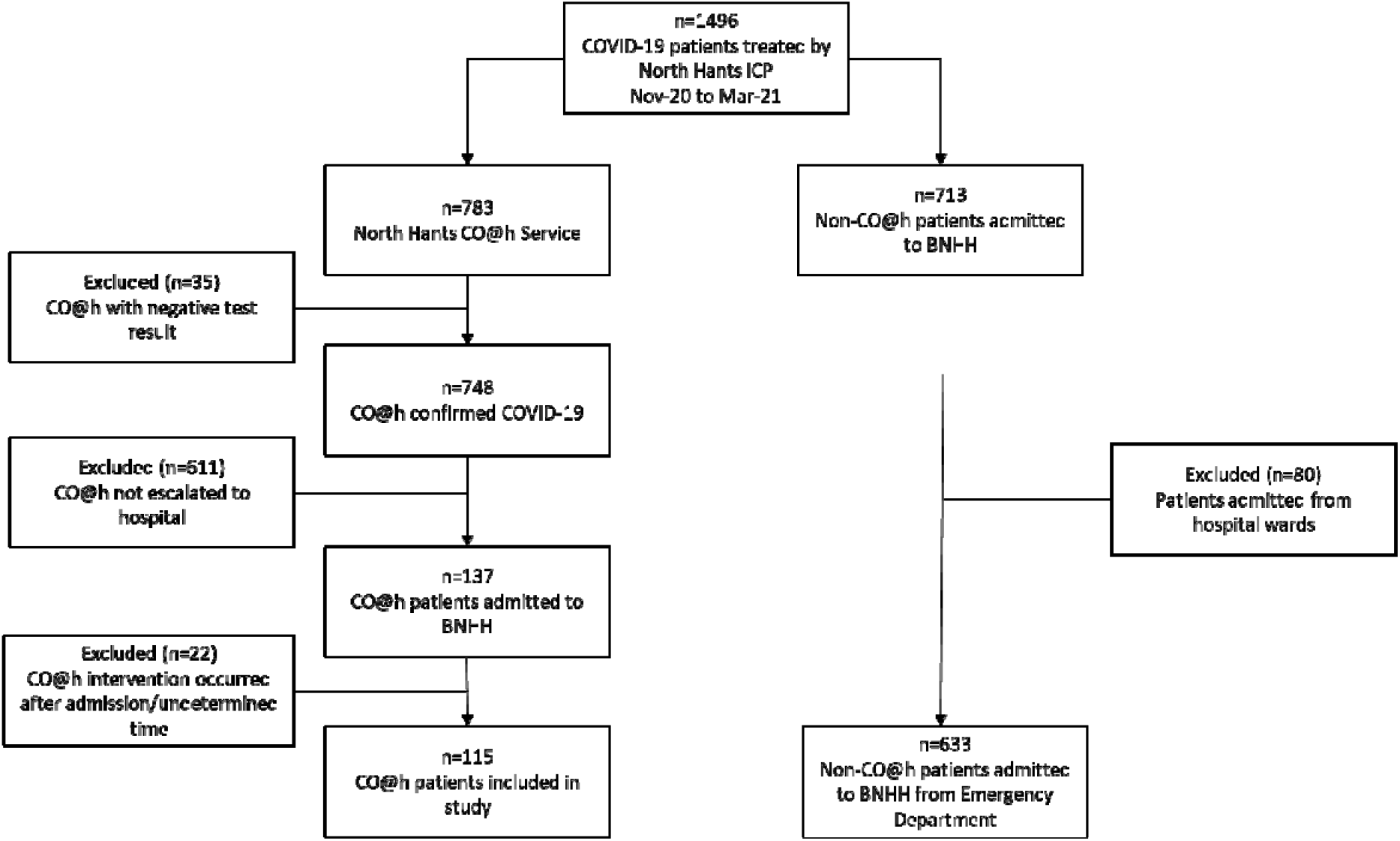
Patient cohort selection showing CO@h intervention group and Non-CO@h control group (those patients not being treated by the CO@h service)

**Figure 3:**
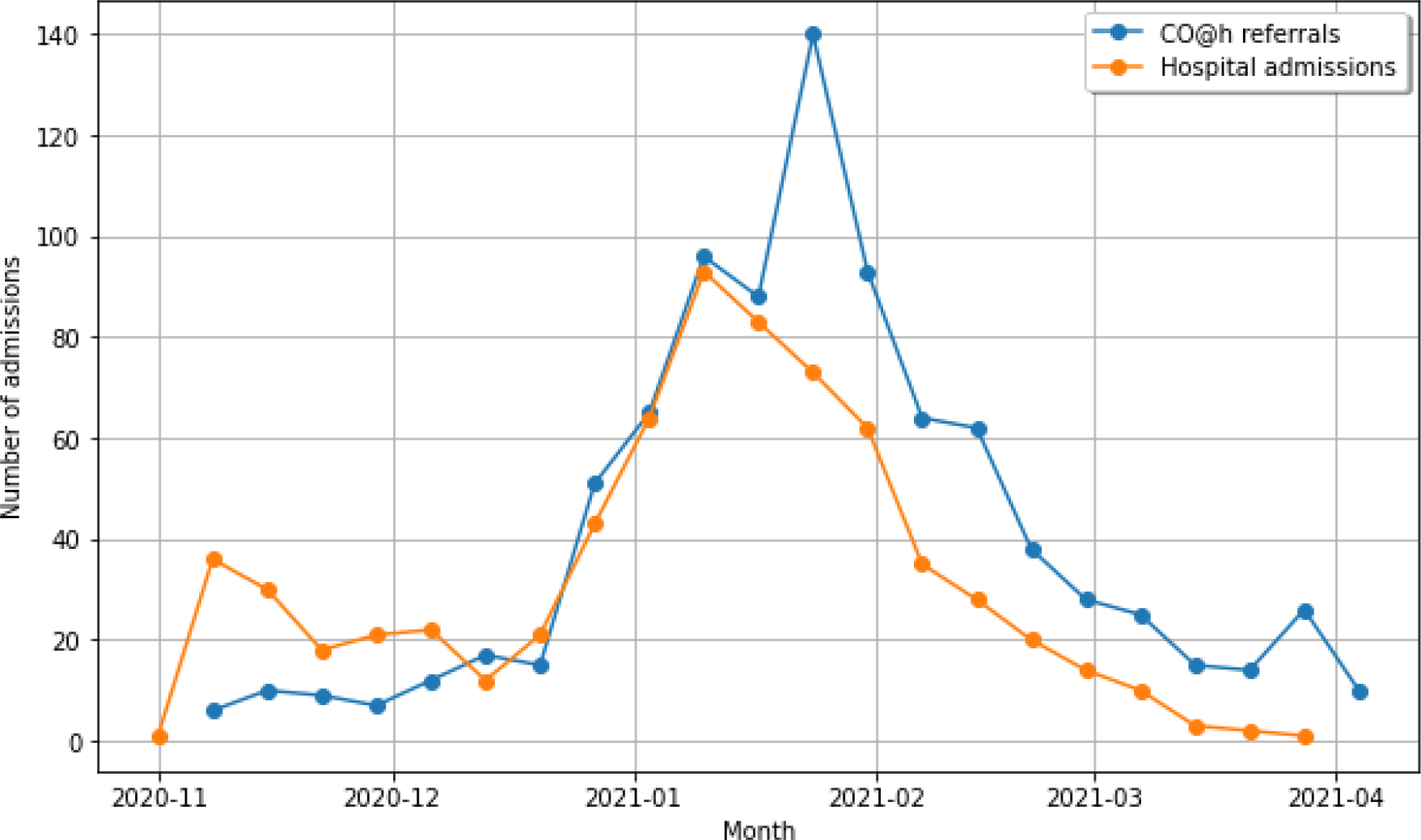
Monthly COVID-19 referrals for monitoring by CO@h service (blue) and total hospital admissions (orange) from November 2020 to March 2021. Hospital admissions include CO@h referrals escalated and admitted to hospital.

Distributions representing the hospital length of stay for both the intervention and control groups are shown in Figure 4. The 80^th^ percentile hospital length of stay for the intervention group was 9 days, 12 days shorter than the 21 days for the control group. The mean hospital length of stay was 7.1 days (95% CI 5.7 -8.5 days) in the intervention group, and 13.2 days (95% CI 12.2 – 14.1 days) in the control group.

**Figure 4:**
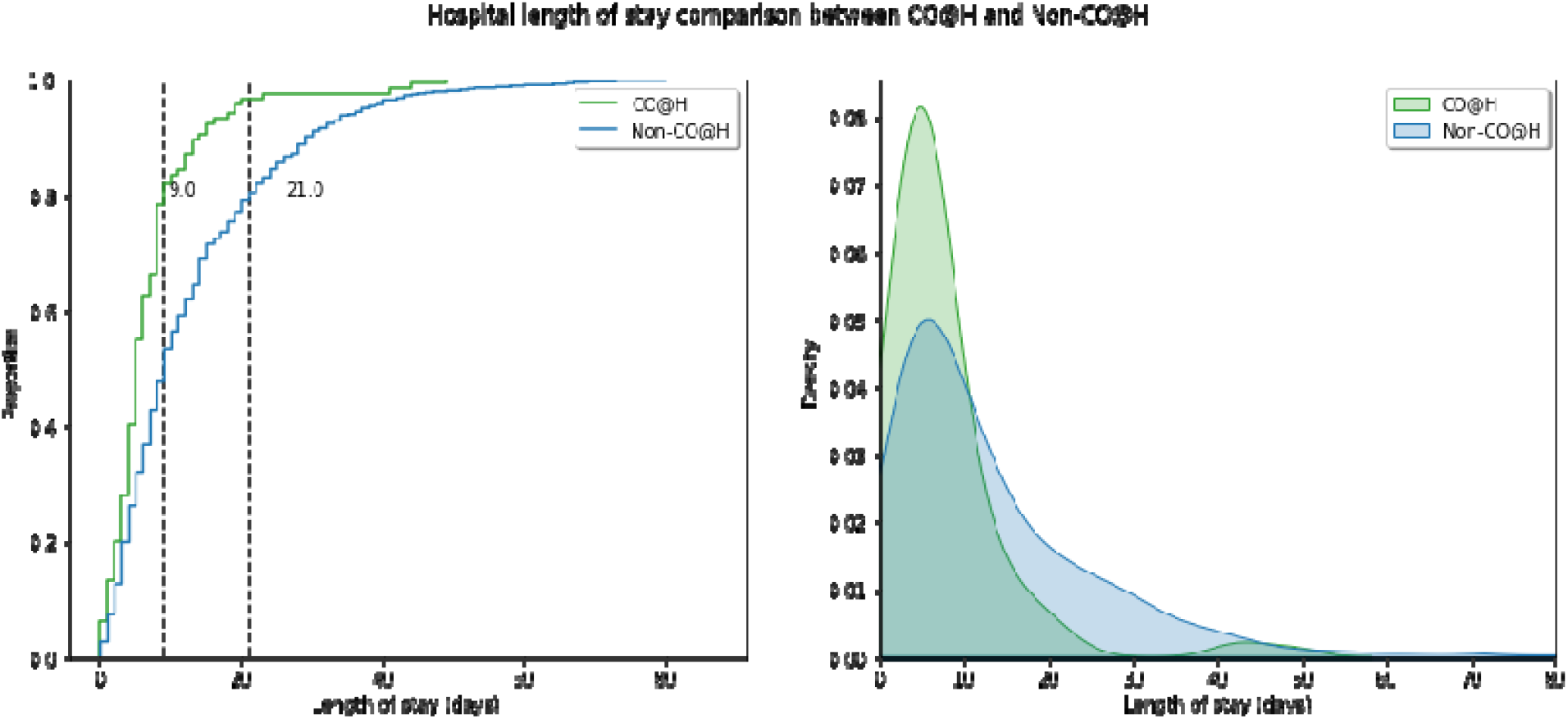
Hospital length of stay distributions for our intervention (CO@h; green) and control cohorts (Non-CO@h; blue). (Left) cumulative distribution for length of stay with 80th percentile length of stay shown as vertical lines for each period. (Right) kernel density estimate for hospital length of stay.

Table 1 gives a contingency table showing the age group, gender, and comorbidity risk for patients in the intervention and control groups ascertained from medical records. Pearson’s Chi-squared test identified that there were no significant differences between the distributions of the age group, gender, and comorbidity risk.

**Table 1.** Contingency table of patients’ age group, gender and comorbidity risk with positive SARS-CoV-2 test result. Brackets give characteristic group percentages.

| Characteristic | CO@h | Non-CO@h |
| --- | --- | --- |
| **No.** | 115 | 633 |
| **Age Group** |  |  |
| 0-19 | 0 (0) | 9 (1.42) |
| 20-45 | 12 (10.43) | 93 (14.69) |
| 46-65 | 43 (37.39) | 167 (26.38) |
| 66-85 | 47 (40.87) | 263 (41.54) |
| 85+ | 13 (11.30) | 101 (15.96) |
| **Gender** |  |  |
| Female | 61 (53.04) | 289 (45.66) |
| Male | 54 (46.96) | 344 (54.34) |
| **Comorbidity Risk** |  |  |
| Yes | 82 (71.30) | 456 (72.04) |
| No | 33 (28.70) | 177 (27.96) |

In Table 2, we summarise the odds ratios and 95% confidence interval for each outcome. Crude odds ratios for CO@h are calculated along with adjusted odds ratios.

**Table 2:** Crude odds ratios for CO@h and odds ratios adjusted for age, gender, and COVID-19 comorbidities in predicting outcomes of Hospital length of stay > 3,7,14 and 28 days, ICU admission, hospital mortality. Significance levels of each adjusted odds ratios variable are given as *** for p<0.001, ** for p<0.01, * for p<0.05 and blank otherwise.

| Variable | Crude OR | Adjusted OR |
| --- | --- | --- |
| **Outcome i: Hospital LOS > 3 Days** |  |  |
| **CO@h \*** | 0.64 (0.41, 1.01) | 0.62 (0.39, 1.00) |
| **Gender** |  |  |
| Female (ref) | - | 1 |
| Male | - | 1.27 (0.88, 1.86) |
| **Age Group** |  |  |
| 0-19 ** | - | 0.05 (0.00, 0.27) |
| 20-45 | - | 0.64 (0.38, 1.08) |
| 46-65 (ref) | - | 1 |
| 66-85 ** | - | 2.10 (1.34, 3.32) |
| 85+ *** | - | 4.66 (2.21, 11.05) |
| **Comorbidity risk** | - | 1.17 (0.75, 1.80) |
| **Outcome ii: Hospital LOS > 7 Days** |  |  |
| **CO@h \*\*\*** | 0.38 (0.25, 0.58) | 0.35 (0.22, 0.54) |
| **Gender** |  |  |
| Female (ref) | - | 1 |
| Male | - | 1.28 (0.93, 1.77) |
| **Age Group** |  |  |
| 0-19 | - | 0.17 (0.01, 0.99) |
| 20-45 *** | - | 0.34 (0.19, 0.60) |
| 46-65 (ref) | - | 1 |
| 66-85 *** | - | 2.30 (1.58, 3.38) |
| 85+ *** | - | 3.78 (2.24, 6.52) |
| **Comorbidity risk \*** | - | 1.50 (1.01, 2.22) |
| **Outcome iii: Hospital LOS > 14 Days** |  |  |
| **CO@h \*\*\*** | 0.23 (0.12, 0.42) | 0.22 (0.11, 0.41) |
| **Gender** |  |  |
| Female (ref) | - | 1 |
| Male | - | 1.07 (0.75, 1.51) |
| **Age Group** |  |  |
| 0-19 | - | - |
| 20-45 | - | 0.44 (0.18, 0.97) |
| 46-65 (ref) | - | 1 |
| 66-85 ** | - | 1.91 (1.23, 2.98) |
| 85+ *** | - | 2.79 (1.65, 4.76) |
| **Comorbidity risk \*\*\*** | - | 2.69 (1.62, 4.61) |
| **Outcome iv: Hospital LOS > 28 Days** |  |  |
| **CO@h \*** | 0.21 (0.05, 0.57) | 0.21 (0.05, 0.59) |
| **Gender** |  |  |
| Female (ref) | - | 1 |
| Male | - | 1.07 (0.65, 1.76) |
| **Age Group** |  |  |
| 0-19 | - | - |
| 20-45 | - | 0.59 (0.13, 1.95) |
| 46-65 (ref) | - | 1 |
| 66-85 | - | 1.67 (0.87, 3.39) |
| 85+ * | - | 2.37 (1.12, 5.17) |
| **Comorbidity risk \*** | - | 3.20 (1.38, 8.76) |
| **Outcome v: ICU admission** |  |  |
| **CO@h** | 0.51 (0.17, 1.19) | 0.43 (0.15, 1.04) |
| **Gender** | - | - |
| Female (ref) | - | 1 |
| Male | - | 1.66 (0.93, 3.03) |
| **Age Group** |  |  |
| 0-19 | - | - |
| 20-45 | - | 0.94 (0.42, 2.01) |
| 46-65 (ref) | - | 1 |
| 66-85 *** | - | 0.21 (0.10, 0.40) |
| 85+ ** | - | 0.04 (0.00, 0.18) |
| **Comorbidity Risk \*\*** | - | 3.15 (1.56, 6.80) |
| **Outcome vi: 30-day hospital mortality** |  |  |
| **CO@h \*\*\*** | 0.21 (0.08, 0.46) | 0.21 (0.08, 0.47) |
| **Gender** |  |  |
| Female (ref) | - | 1 |
| Male * | - | 1.53 (1.02, 2.31) |
| **Age Group** |  |  |
| 0-19 | - | - |
| 20-45 | - | - |
| 46-65 (ref) | - | 1 |
| 66-85 *** | - | 4.80 (2.62, 9.43) |
| 85+ *** | - | 7.32 (3.71, 1.53) |
| **Comorbidity Risk** | - | 1.95 (1.03, 3.96) |

Adjusted odds ratios for CO@h show an association with a reduction for several adverse patient outcomes: 30-day hospital mortality (p<0.001 OR 0.21 95% CI 0.08-0.47), hospital length of stay larger than 3 days (p<0.05, OR 0.62 95% CI 0.39-1.00), 7 days (p<0.001 OR 0.35 95% CI 0.22-0.54), 14 days (p<0.001 OR 0.22 95% CI 0.11-0.41), and 28 days (p<0.05 OR 0.21 95% CI 0.05-0.59). No significant reduction ICU admission was observed (p>0.05 OR 0.43 95% CI 0.15-1.04).

Within the CO@h cohort 6/115 (5.22%) had a 30-day mortality outcome compared with 130/633 (20.54%) for Non-CO@h demonstrating a 25.40% reduction. The CO@h cohort included 5/115 (4.35%) that were admitted to ICU in comparison to 52/633 (8.21%) for Non-CO@h giving a 52.93% reduction. No patients were readmitted to hospital within 30 days of hospital admission for those admitted from CO@h, compared with 55/633 (8.7%) for Non-CO@h.

## Discussion

The CO@h initiative was implemented nationally to protect patients by improving early recognition of deterioration in COVID-19 and to protect the healthcare system from being overwhelmed with inappropriate admissions. The CO@h service was universally implemented across England (by Feb 2021) over a period of 3 months.

In this QI initiative we focussed on evaluating the efficacy of CO@h by retrospectively evaluating COVID-19 patient outcomes for those admitted to CO@h in North Hampshire Primary Care Network and subsequently being admitted to hospital. The initiative achieved its aim with reductions in length of stay and 30-day mortality rates for patients admitted to hospital via the CO@h pathway relative to direct hospital admission. In the majority of acute COVID-19 cases, severe hypoxia (often presenting ‘silently’ without breathlessness) has been a significant contributor in patient deterioration from pneumonia to acute respiratory distress syndrome (Couzin-Frankel, 2020) (Greenhalgh, et al., 2021) (Brouqui, et al., 2021) (Knight & al., 2020) (O’Carroll, et al., 2020). Pathologically, hypoxia due to COVID-19 is likely driven by a mixture of intrapulmonary shunting, instability in lung perfusion, intravascular microthrombi, and alveolar collapse (Couzin-Frankel, 2020) (Levitan, 2020). In particular, silent hypoxia from COVID-19 is associated with rapid deterioration (i.e. patients immediately being admitted to ICU from home or shorter ward stays) and higher rates of respiratory failure (Brouqui, et al., 2021). Driving factors include unrecognised lung function decline leading to damage to the brain and central nervous system (Rahman, et al., 2021). Early identification and proactive management of severe COVID-19, directly improves patient outcomes (O’Carroll, et al., 2020) (Brouqui, et al., 2021) (Vindrola-Padros, et al., 2020). Prospective outcomes can be improved by prompt prescription of medication such as Dexamethasone (The RECOVERY Collaborative Group, 2021) and trials of non-invasive ventilation (Menzella, et al., 2021). To our knowledge, this QI service evaluation is the first to demonstrate that CO@h is associated with improved patient outcomes for an NHS trust (i.e., mortality, ICU admission and length of stay). This CO@h service was implemented as part of the national framework (NHS England, 2020) and therefore these findings should be of interest to future CO@h operations in response to the pandemic. CO@h services have now been provisioned internationally, for example, StepOne have applied the CO@h model to support intervention of COVID patients across 16 states in India (George Institute, 2021).

COVID-19 is endemic worldwide, and hence, there is an urgent need for optimised early identification of patient deterioration for patients at home. As healthcare systems aim to restore elective activity, the backlog of non-COVID patients requiring intervention is stark. In England, the British Medical Association estimates there were 3.37 million fewer elective procedures and 21.4 million fewer outpatient attendances between April 2020 and March 2021 (Association, 2021). The COVID-19 pandemic has generated research into effective and streamlined patient care which has ramifications beyond the context of the pandemic. CO@h is one aspect of the nationally led programme NHS @home; which aims to maximise the use of technology to support more people to better self-manage their health and care at home (NHS England, NHS @home, 2021). With access to more timely preventative care, patient burden on both primary and emergency care can be reduced while providing patients more personalised intervention. In particular, home pulse oximetry has long been used in primary care settings as a cost-effective approach to monitor chronic lung conditions and heart disease (Plüddemann, Thompson, Heneghan, & Price, 2011), and there is a growing evidence base for the model’s effectiveness and safety in COVID-19 (Vindrola-Padros, et al., 2020) (Inada-Kim, et al., 2020) (Boniface, Zlatev, Guerrero-Luduena, & Armitage, 2020) (Greenhalgh, et al., 2021) (Nunan, et al., 2020). Further prospective studies are required to understand how remote monitoring can be implemented in wider contexts, potentially focussed on high-risk patients with significant co-morbidities.

These findings must be understood in light of their limitations. CO@h was rapidly developed in response to the pandemic, and as a result, the improvement cycles were conducted at pace. PDSA quality improvement was conducted using evidence-based practice, where insights were provided by data professionals to clinicians. A multi-disciplinary team of healthcare professionals, QI personnel, and data scientists met frequently to discuss patient care, and CO@h efficacy. Operational improvements were implemented through these discussions to deliver continual improvement especially procedures relating to integrated services between conveyance, CO@h and hospitals. Formally, the distinct improvement cycles were as follows: (1) CO@h service pilots (1^st^ wave of the pandemic: March 2020 to July 2020) including community COVID-19 assessment centres implemented without remote monitoring beyond paper diaries and phone, treating n=1600 suspected-COVID patients and escalating n=105 to hospital; and support by hospital Same Day Emergency Care and care homes telemedicine services (2) NHS Trust-wide implementation of CO@h (2^nd^ wave of the pandemic: November 2020 to March 2021) with efficacy evaluation presented here.

Although we show that patients admitted to hospital after CO@h had better outcomes relative to an unmonitored comparison group, the suggestion that this was driven by CO@h is complicated by other factors. For example, the severity of illness at the time of admission to hospital is not considered in our study. Patient severity from CO@h referrals are likely to be within red and amber categories (in accordance with standard operating procedure), physiological measurement data for all patients at the time of hospital attendance was not available for analysis. This limits inferences that can be made regarding the timely escalation of patients to hospital between each cohort, while many may have stayed at home and survived had their SpO_2_ not been monitored. A related limitation, but likely to be a minor factor in this study, is that measurements are self-reported by patients rather than taken by a healthcare professional. This may introduce some inaccuracy in measurements used for CO@h escalation decisions. Our study did not have a mode of assessing mortality outside of the hospital and therefore may be under reporting mortality for patients in both cohorts. Although, we have considered objective differences regarding age, gender and comorbidities, there may be other qualitative socio-economic or demographic differences that may influence the outcomes not considered in our study. Finally, this service evaluation is for an integrated community care pathway and a single hospital trust, therefore generalisation is limited by population size and clinical setting.

## Conclusion

Our study has shown that COVID Oximetry @home (CO@h) has a significant association with better patient outcomes. To our knowledge, this is the first QI report concerning the efficacy of CO@h service and evaluation of hospital mortality, ICU admission, and length of stay benefits for an NHS Hospital Trust. COVID-19 patients admitted to the CO@h service have been found to have significantly reduced odds of longer length hospital stays, ICU admission and of hospital mortality. These findings demonstrate that, despite the study limitations, CO@h should be considered nationally and internationally in response to the ongoing pandemic and that larger evaluations of efficacy and quality should be undertaken.

## Acknowledgements

We thank Claire Parker from Hampshire, Southampton & Isle of Wight Clinical Commissioning Group for support during the specification, delivery, and evaluation of CO@h. We extend our special thanks to the CO@h Clinical Team who cared for patients and collated data including Alison Hullah (Lead Advanced Nurse Practitioner), Roisin Howell (Lead Advanced Nurse Practitioner) and Giselle Beaumont (Care Coordinator). This report received funding from the NHSX RECOxCARE (Remote oximetry in community care for COVID-19 patients) project and from NHS England to support data collection. The views expressed in this publication are those of the author(s) and not necessarily those of the funders.

